# Sustaining Engagement in HIV Prevention Program Among Adolescents and Young Adults (AYA) in Nigeria: Longitudinal Secondary Analysis

**DOI:** 10.64898/2026.09.03.26360266

**Authors:** Abdulhammed Opeyemi Babatunde, Titilola Gbaja-Biamila, Weiming Tang, Suzanne Day, Susan Nkengasong, Ucheoma Nwaozuru, Nora E. Rosenberg, Yusuf Babatunde, Onyeama Frances, Conserve Donaldson, Tahlil Kadija, Temitope Ojo, Olusanya Olufunto, Chinyere Arinze, Peter Kalulu, Lateef Akeem, David Oladele, Adesola Z. Musa, Folahanmi Akinsolu, Tomilola Musari-Martins, Kathryn Muessig, Rhonda BeLue, Hong Xian, Gbenga Ogedegbe, Collins O. Airhihenbuwa, Juliet Iwelunmor, Oliver Ezechi, Joseph D. Tucker

## Abstract

**Background:** Adolescent and young adult (AYA; 14-24 years old) engagement in HIV programs is often transient, and socioeconomically marginalized AYA are less engaged. Few studies have examined whether engagement can be sustained over time in HIV prevention programs. This study examines correlates of sustained AYA engagement over 24 months in an HIV prevention program in Nigeria.

**Methods:** We conducted a secondary data analysis of longitudinal data from an HIV prevention program among AYA in 32 local communities in Nigeria from 2021 to 2023. In the study, participants were invited to join crowdsourcing open calls, peer-led support meetings, and other activities. Engagement was assessed using a modified 12-item Tiffany-Eckenrode Program Participation Scale (0-4 per item) at months 3, 6, 12, 18, and 24. Sustained AYA engagement was defined as a mean score ≥3.0/4.0 in at least three consecutive contacts (approximately 12 months). Longitudinal analysis was conducted using generalized estimating equations, and state-level contextual analysis was done using a linear mixed model.

**Results:** A total of 1,259 AYA were included. The mean age was 19.8 ± 2.7 years; most were students (n=954,75.8%) and unemployed (n=810,64.3%). The mean participation score remained consistently high throughout the 24 months (3.245 ±0.535 to 3.319 ±0.556), with 73.0% demonstrating sustained engagement. Students were less likely to sustain engagement compared to non-students (aRR = 0.91; 95% CI: 0.84-0.97). Unemployed AYA were more likely to sustain engagement than employed AYA (aRR= 1.09; 95% CI:1.03-1.14). States with higher poverty had significantly greater sustained AYA engagement. All the main findings were robust in sensitivity analyses.

**Conclusion:** AYA engagement in the HIV self-testing implementation was high and sustained over 24 months, especially among socio-economically disadvantaged populations. This study supports the use of a participatory, AYA-led approach in sustaining AYA engagement in HIV prevention programs and has implications for the equity of HIV programs.

## Introduction

Sustaining adolescents and young people (AYA, ages 14-24) engagement in health programs is a major challenge for global health implementation. This is compounded by the current disruption in global health funding, especially in HIV programs in African countries.^1^ Africa accounted for half of the new HIV cases in 2024, and AYA were disproportionately affected.^2,3^ HIV prevention programs in Africa struggle to sustain AYA engagement beyond initial contact, limiting the potential impact.^4^ Nigeria has the largest AYA population and the third highest number of people living with HIV in any African country.^5^ Access to HIV prevention programs among AYA in Nigeria is suboptimal.^6^ Available prevention programs are not tailored to AYA, resulting in social stigma and discrimination.^7^ With the decline in external funding for HIV programs, sustainability of HIV prevention programs among AYA in Nigeria is threatened.

Despite WHO/UNICEF recommendations for meaningful AYA engagement in HIV programs, most HIV research in Africa still excludes AYA from study design and implementation. A scoping review found that 88% of the 74 interventions on HIV prevention among AYA in Africa had minimal or no youth engagement.^4^ In addition, the duration of reported engagement is often unclear and short.^4,8,9^ Unstable AYA engagement might reverse the initial gains of the intervention, limiting the potential effect of interventions that aimed to improve HIV testing, adherence to Pre-Exposure Prophylaxis (PrEP), and linkage to care.^10,11^ Sustained AYA engagement can promote community trust, willingness to engage in future research, and inclusivity of marginalized groups.^12^ Sustaining AYA engagement in HIV research in Nigeria is crucial for achieving the 2030 target to end AIDS as a public health concern.^13^

Available evidence suggests that social determinants of health, such as poverty and unemployment, contribute to low engagement of AYA in HIV care in Africa.^3^ This threatens health equity among marginalized AYA in accessing HIV prevention programs. Participatory community-based approaches, such as crowdsourcing open calls, have been shown to promote the inclusion of poor, unemployed and marginalized AYA with lived experience in HIV prevention programs.^9,14^ This may strengthen ownership and AYA engagement. However, there is limited evidence on sustaining AYA engagement through a participatory approach.

The Innovative Tools for Expanding HIV Self Testing (ITEST) was an implementation research study that engaged AYA using a series of participatory activities to promote HIV self-testing, PrEP uptake, and linkage to care among AYA in Nigeria.^15,16^ Participatory activities included crowdsourcing open calls, peer-led support meetings, and AYA research assistants. This study examines correlates of sustained AYA engagement in the ITEST HIV prevention program over 24 months in Nigeria.

## Methods

### Study design

This is a secondary data analysis of data collected during the pragmatic stepped-wedge cluster randomized controlled trial (RCT) “A Pragmatic Trial to Expand Youth-friendly HIV Self-testing (I-TEST)” (NCT04710784) from April 2021 to August 2023.^15^ “This secondary analysis follows a longitudinal cohort design and is reported using the STROBE checklist.^17^

### Study Participants

Study participants were aged 14-24 years, residing in any of the selected 32 local government areas (LGAs) across 14 states in Nigeria, and owned or had access to a mobile phone. Potential participants were screened and recruited by trained AYA research assistants through AYA-centered activities. AYA who reported illness, cognitive impairment, living with HIV, or tested positive at screening were excluded. AYA, who would not be residing in the same location for the next two years were also excluded.^15^ A sample size of 1,248 was determined by the parent RCT which also provided adequate power for the longitudinal analysis.^18^

### AYA Engagement and Public Involvement

Participants were engaged in designing the intervention and during the intervention through AYA-led activities such as crowdsourcing open calls (a group of people attempts to solve all or part of a problem, then shares solutions with the public),^19^ peer-led support meetings, and AYA research assistants (Figure 1).

**Figure 1:**
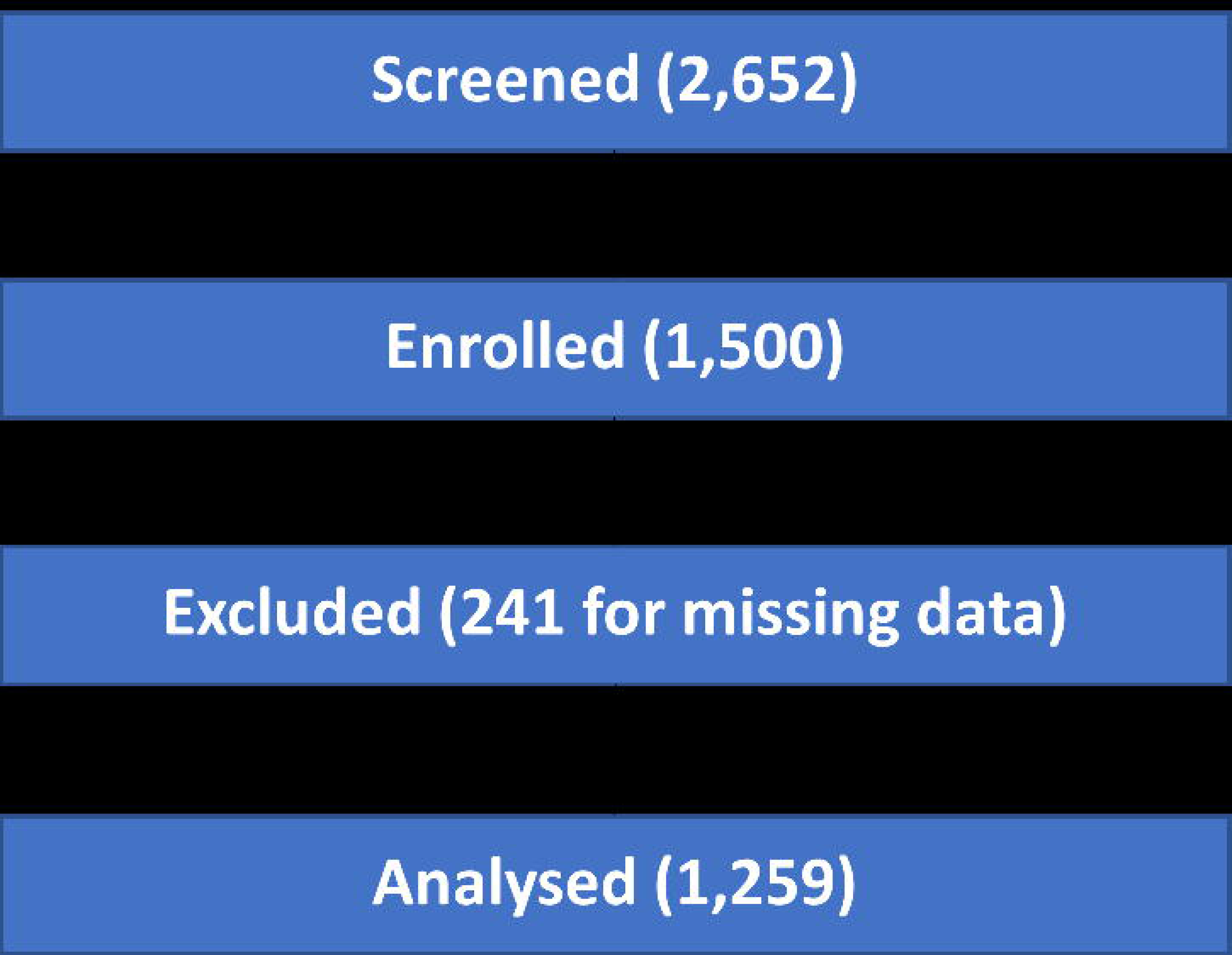
Youth Participatory Action Research Activities in the ITEST HIV Prevention program. Figure shows participatory activities for AYA engagement from conceptualization to implementation and sustaining ITEST HIV prevention across 32 LGAs and 14 states in Nigeria.

### Variables and instruments

Variables extracted from the baseline RCT dataset included socio-demographic information (age, sex, ethnic group, level of education, employment status, marital status, income, local government area), sexual behavior history, and AYA engagement at months 3 (baseline), 6, 12, 18, and 24.

AYA engagement was assessed using the 12-item modified Tiffany-Eckenrode Program Participation Scale. This scale has four subscales relating to engagement in HIV prevention programs: youth ownership (six items), voice/influence (three items), support (one item), and involvement (two items). The scale is a reliable measure of the quality of AYA program engagement ^20^ and has been used among Nigerian AYA.^16^ The survey assessed AYA engagement over the preceding three months based on self-report at each time point.

AYA engagement was assessed using a Likert scale, which was graded 0 to 4 based on the level of agreement with each statement (strongly disagree, disagree, neutral, agree, and strongly agree) across 12 items. A mean participation score of greater than or equal to 3.0/4.0 was categorized as engaged. This is based on the interpretation of the grading in a previous similar publication assessing AYA participation in a pilot study.^16^ AYA engagement was considered sustained by maintaining a mean score greater than or equal to 3.0/4.0 at three consecutive contacts (approximately 12 months) to align with an existing definition.^21^

### Sustainability Framework

We used the Dynamic Sustainability Framework as a theoretical basis for the analysis.^22^ The framework captures how engagement is an iterative process and sustained engagement is influenced by individual, programmatic, and contextual correlates. For this study, sustained AYA engagement was defined as maintaining AYA participation in the HIV prevention program for 12 or more months.^23,24^

### Data analysis

Statistical analyses were carried out using Microsoft Excel and SPSS 26. Efforts to minimize bias in the primary trial including standardization of data collection and youth research assistant training were described in published study protocol.^18^ For this secondary analysis, potential selection bias from missing data was addressed in two ways. Participants with no responses to engagement scale at any time point were excluded from analysis (complete case analysis) while participants with partial item-level missing data within a time point were completed with mean score of available items. We conducted sensitivity analysis comparing the sociodemographic characteristics of included and excluded data. Descriptive analysis of the baseline sociodemographic characteristics was presented using frequency tables and charts. The mean, SD, and range of engagement scores were calculated across all 12 items. Mean engagement scores were used to dichotomize each participant into engaged (≥3.0/4.0) or unengaged (<3.0/4.0). Longitudinal analysis was conducted using a generalized estimating equations (GEE) model to estimate the repeated-measures predictive relationship between the engaged and unengaged participants. These equations accounted for the within-subject correlation of repeated measures of engagement across all follow-ups. The bivariate analysis with a log link function estimated the adjusted relative risk (aRR) of engagement, accounting for time point and other independent variables such as sex, age, LGA, student and employment status. A state-level contextual analysis of GDP per capita,^25^ literacy rate,^26^ poverty rate,^27^, and HIV prevalence^28^ was done using a linear mixed model. We used the most recent available data before 2023, when the RCT ended. This showed the longitudinal interaction at the clustered level that correlates with AYA sustainment. We used state-level data because this has greater relevance for HIV policy in Nigeria. We also conducted a comparative analysis using a chi-square test to test for association between socio-demographic characteristics and sustained AYA engagement. A p-value of 0.05 was considered significant.

### Ethical Considerations

Ethical approval for the RCT was granted by the Nigerian Health Research Ethics Committee (NHREC) and the Nigerian Institute of Medical Research Institutional Ethical Review Boards IRB/18/028. The procedure for data collection and management complied with ethical standards and aligned with the Helsinki Declaration. All the subjects signed informed consent, and data were anonymized using a unique ID.

## Results

A total of 1259 of 1500 participants were included in the study (Figure 2). The mean age was 19.8 (SD = 2.67). Most participants (728, 57.8%) were aged 20-24. A total of 641 (50.9%) were girls or women, and most were Igbo tribe (476, 37.8%), followed by Yoruba (352, 27.9%). There were 23-61 participants from each of the 32 LGAs spanning four geopolitical zones in the country, mostly south-south (475, 37.7%). Most AYA (845, 67.1%) completed secondary education, and 954 (75.8%) were students. Only 166 (13.2%) were employed. A total of 1232 (97.9%) participants were never married or single, and most (561, 44.6%) earned greater than the minimum wage of $558 annually (exchange rate at beginning of trial, N387 = $1) (Table 1). At month 3, 74.3% (935) of participants were engaged. This increased to 81.5% (1026) at month 24.

**Figure 2:**
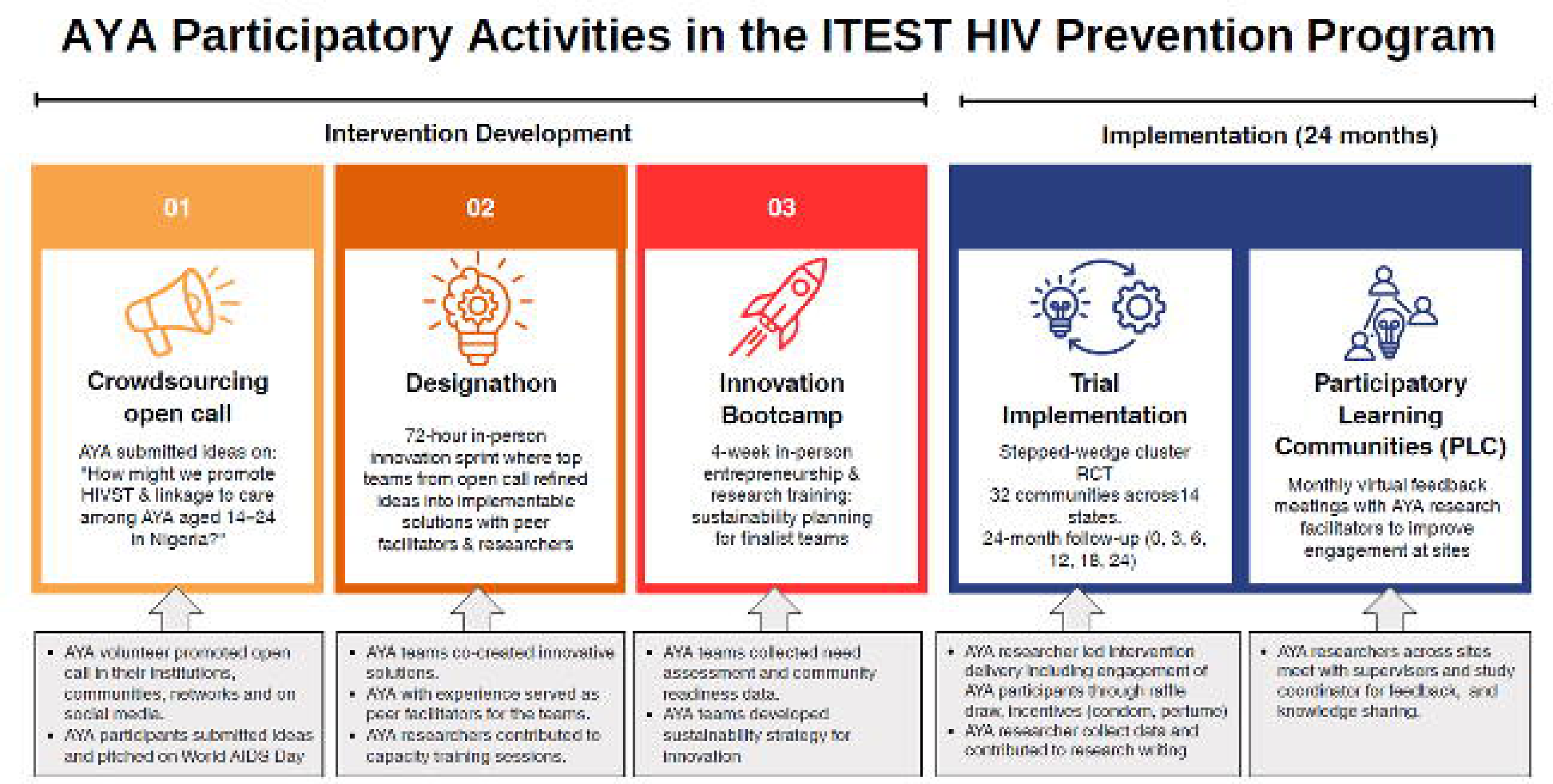
Participant Flow Diagram. Figure showing the number of participants included in the secondary analysis

**Table 1:** Socio-demographic characteristics and sexual history of AYA participants.

|  | **Freq N=1259** | **%** |
| --- | --- | --- |
| **Age (years)** |  |  |
| 19 | 531 | 42.2 |
| ≥20 | 728 | 57.8 |
| **Sex** |  |  |
| Male | 618 | 49.1 |
| Female | 641 | 50.9 |
| **Ethnicity** |  |  |
| Yoruba | 351 | 27.9 |
| Hausa | 41 | 3.3 |
| Igbo | 476 | 37.8 |
| Ibibio | 110 | 8.7 |
| Others | 281 | 22.3 |
| **Geopolitical Zone** |  |  |
| South South | 475 | 37.7 |
| South West | 398 | 31.6 |
| South East | 213 | 16.9 |
| North Central | 173 | 13.7 |
| **Level of education completed** |  |  |
| No Formal | 1 | 0.1 |
| Primary | 157 | 12.5 |
| Secondary | 845 | 67.1 |
| Tertiary | 256 | 20.3 |
| **Student** |  |  |
| Yes | 954 | 75.8 |
| No | 305 | 24.2 |
| **Employment status** |  |  |
| Employed | 166 | 13.2 |
| Self Employed | 283 | 22.5 |
| Unemployed | 810 | 64.3 |
| **Marital Status** |  |  |
| Never Married or Single | 1232 | 97.9 |
| Unmarried Couple Living Together | 16 | 1.2 |
| Married |  |  |
| Divorced or separated | 8 | 0.6 |
| Widowed | 2 | 0.2 |
|  | 2 | 0.2 |
| **Average Annual Income** |  |  |
| <\$558 | 561 | 44.6 |
| ≥\$558 | 276 | 21.9 |
| Do not want to answer | 422 | 33.5 |
| **Disabilities or illness\*\*** |  |  |
| Yes | 49 | 3.9 |
| No | 1148 | 91.2 |
| I don't know | 62 | 4.9 |
| **Ever Had Sex** |  |  |
| Yes | 556 | 44.2 |
| No | 703 | 55.8 |
| **Sexual Relations** |  |  |
| With men only | 223 | 17.7 |
| With women only | 286 | 22.7 |
| With men and women | 6 | 0.5 |
| I don't want to talk about it | 38 | 3.0 |
| **Condom Use (n=297)\*** |  |  |
| Always | 79 | 26.6 |
| Often | 51 | 17.2 |
| Occasionally | 61 | 20.5 |
| Never | 106 | 35.7 |

\*field was optional. only participants who have had sexual intercourse could report condom use

The mean engagement score remained consistently high throughout the 24 months (3.25±0.54 to 3.32±0.56), with 73.0% of participants demonstrating sustained engagement. Perception of having a voice/power to influence decisions in the program had the lowest average score (mean = 3.15, SD 0.09, range = 2.99-3.22). Respectfulness of youth organizers had the highest average score (mean = 3.44, range = 3.40-3.46) (Table 2). Sustained engagement was significantly associated with lower educational level (RR = 1.101, 95% CI 1.101-1.004), not being a student (RR = 1.188, 95% CI 1.113 - 1.269), and previous sexual experience (RR = 0.903, 95% CI 0.845-0.965) (Table 3).

**Table 2:** Average score of participants per contact period for each of 12 item Tiffany-Eckenrode Program Participation Scale.

| Questions | Month 3 Mean<br>(SD) | Month 6 Mean<br>(SD) | Month 12 Mean<br>(SD) | Month 18 Mean<br>(SD) | Month 24 Mean<br>(SD) |
| --- | --- | --- | --- | --- | --- |
| Program activities are interesting | 3.27 (0.66) | 3.30 (0.65) | 3.25 (0.70) | 3.19 (0.75) | 3.17 (0.77) |
| Participating helped to continue to know HIV status | 3.31 (0.68) | 3.38 (0.62) | 3.34 (0.65) | 3.35 (0.65) | 3.36 (0.63) |
| I learnt a lot from participating | 3.23 (0.69) | 3.36 (0.66) | 3.35 (0.69) | 3.34 (0.67) | 3.33 (0.66) |
| The program helped me to take ownership of my health | 3.27 (0.69) | 3.38 (0.65) | 3.35 (0.71) | 3.31 (0.69) | 3.32 (0.67) |
| Youth organizers were respectful | 3.40 (0.65) | 3.46 (0.61) | 3.45 (0.64) | 3.41 (0.65) | 3.45 (0.65) |
| Youth organizers listened to participant | 3.36 (0.67) | 3.42 (0.62) | 3.37 (0.66) | 3.39 (0.68) | 3.36 (0.65) |
| Participants feel they have voice/power to influence decision | 2.99 (0.84) | 3.17 (0.82) | 3.18 (0.80) | 3.21 (0.81) | 3.22 (0.78) |
| Easy to get involved in the program | 3.23 (0.72) | 3.32 (0.69) | 3.31 (0.73) | 3.34 (0.72) | 3.40 (0.69) |
| I felt safe when involved in the program | 3.16 (0.77) | 3.29 (0.70) | 3.28 (0.75) | 3.30 (0.72) | 3.30 (0.70) |
| Youth organizers available to support or help with a problem | 3.27 (0.76) | 3.35 (0.69) | 3.31 (0.77) | 3.35 (0.72) | 3.22 (0.79) |
| Influence how community treat young people | 3.16 (0.76) | 3.27 (0.72) | 3.24 (0.75) | 3.32 (0.75) | 3.25 (0.73) |
| Program involved young people in the community | 3.28 (0.74) | 3.39 (0.65) | 3.35 (0.69) | 3.31 (0.73) | 3.43 (0.68) |

**Table 3.**
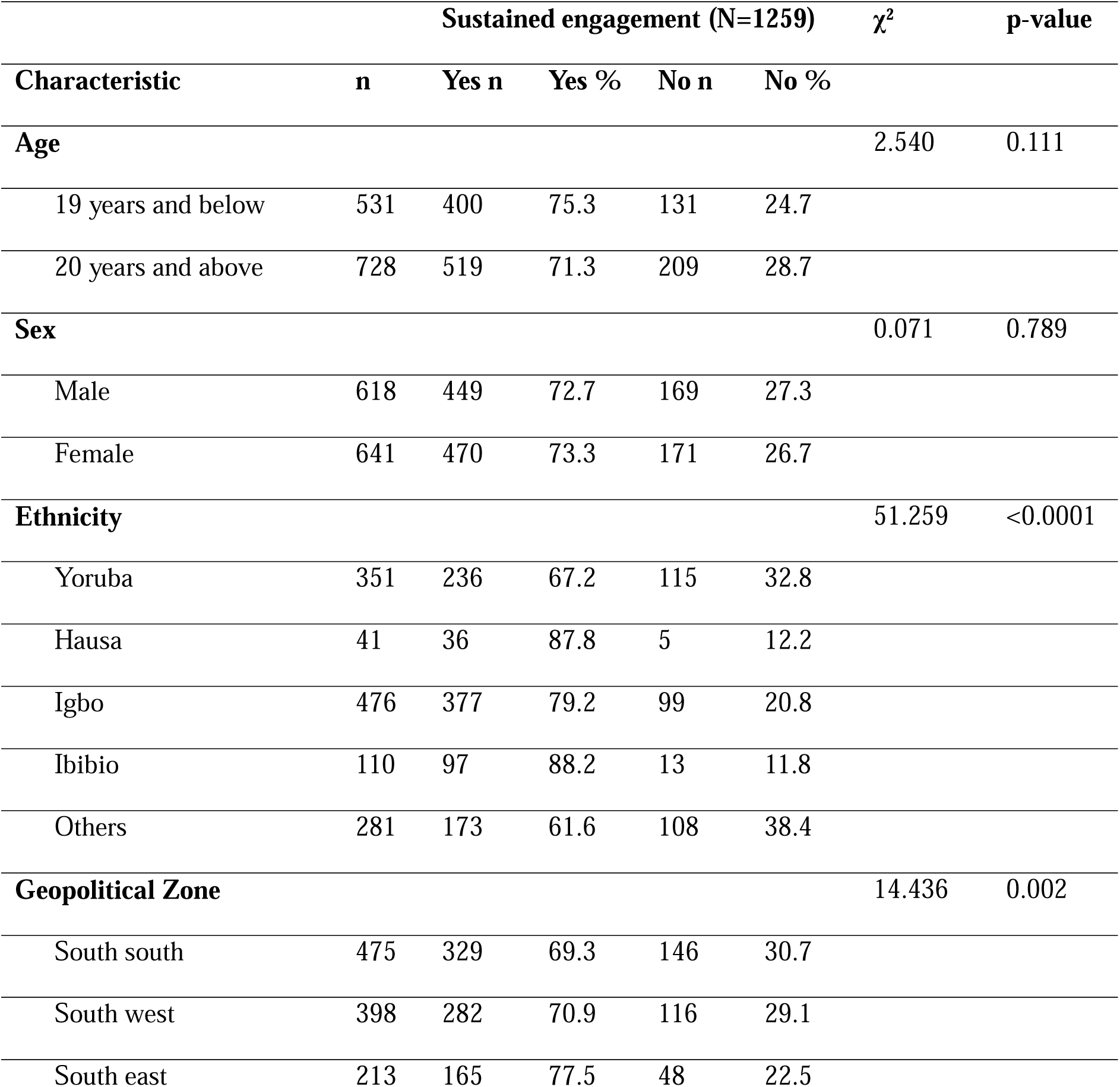
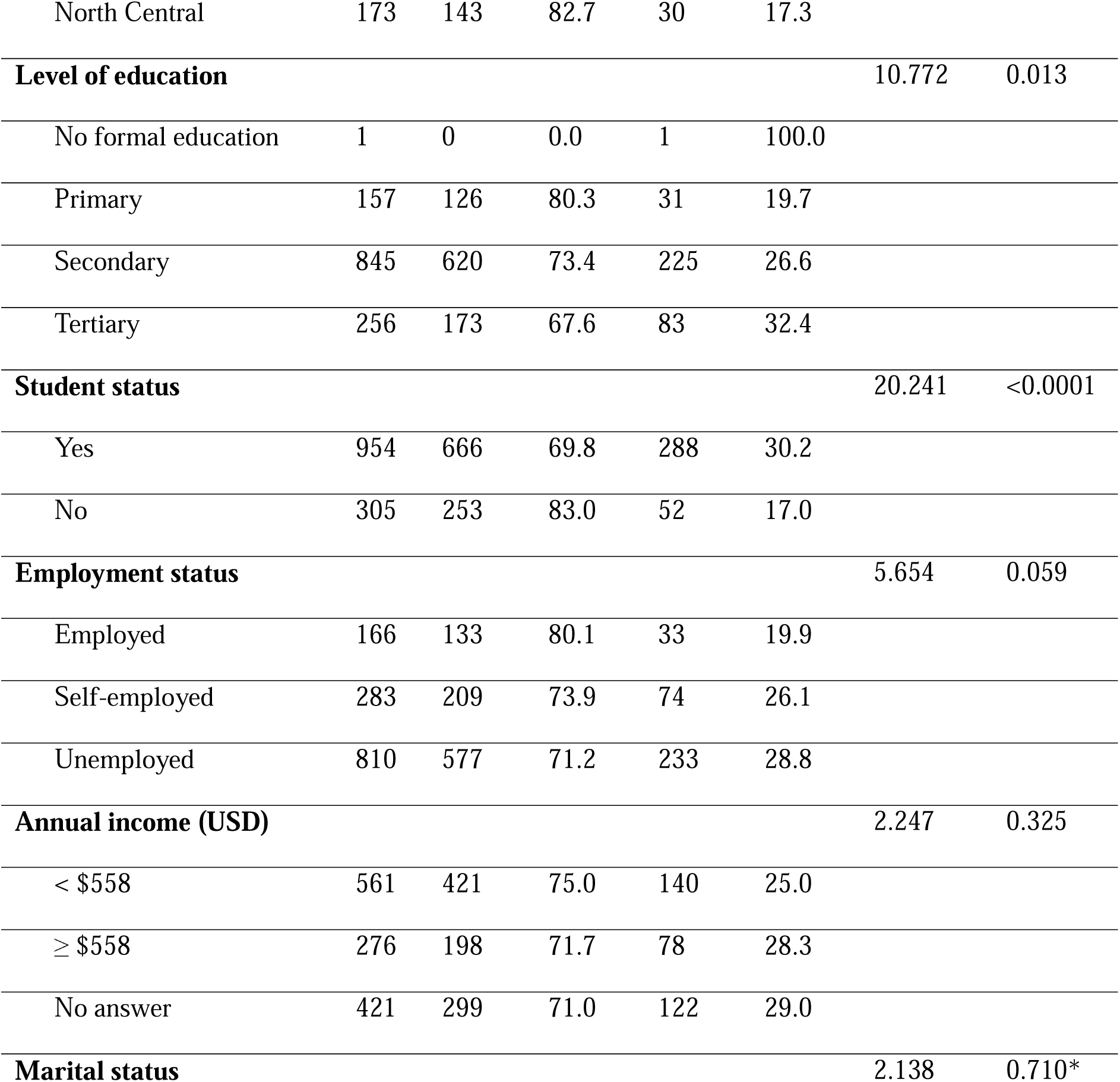
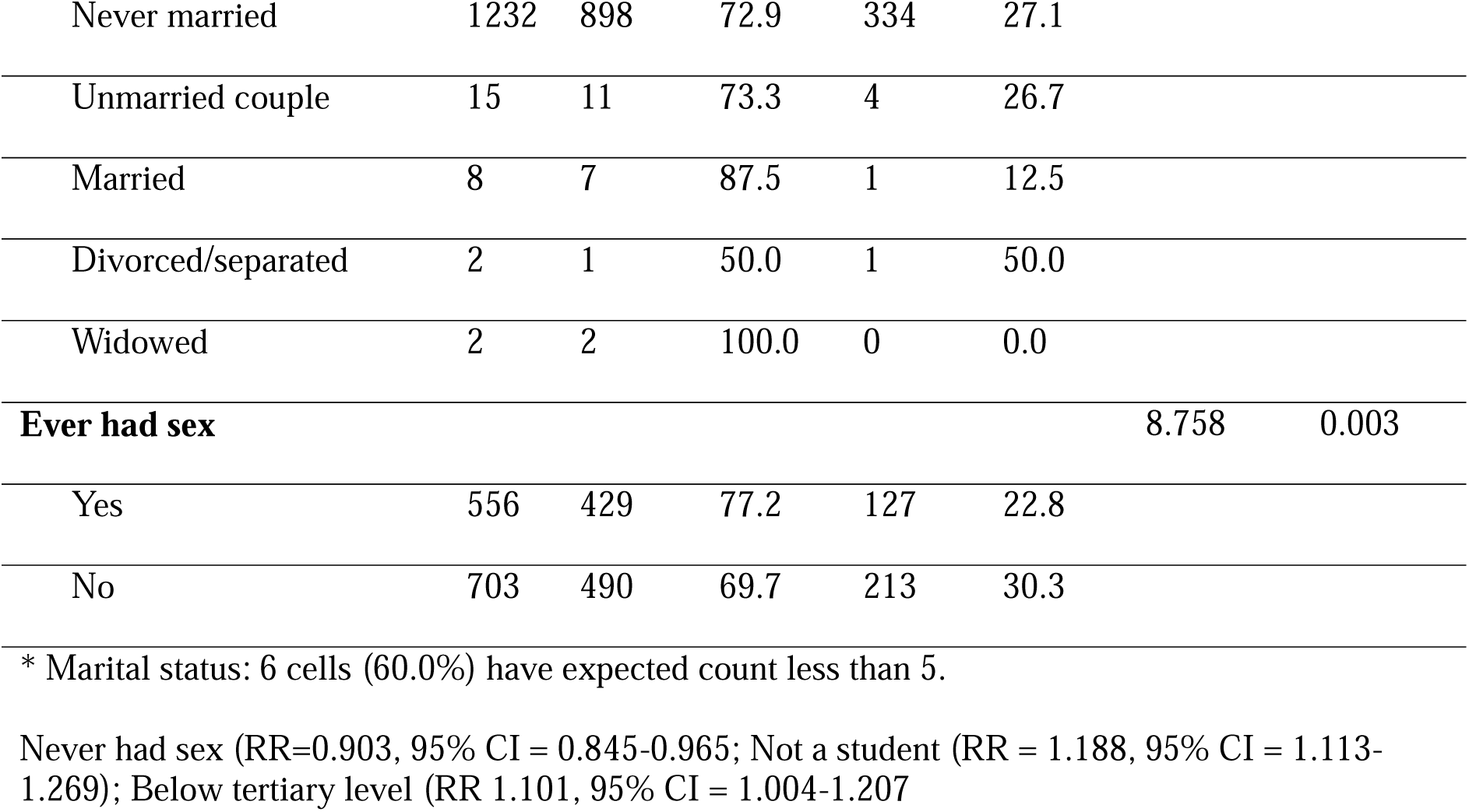
Relationship between sustained youth engagement and socio-demographic characteristics.

GEE analysis showed that compared to engagement at month 3 (baseline), participants were 15% more likely to be engaged at month 24 (adjusted RR (aRR) = 1.15; 95% CI: 1.10–1.18). Students were 9% less likely to be engaged compared to non-students (RR = 0.91; 95% CI: 0.84–0.97). Also, unemployed AYA were 9% more likely to sustain engagement compared to employed (aRR = 1.09; 95% CI: 1.03–1.14). (Table 4). In addition, Cramér’s V analysis showed a statistically significant but weak-to-moderate association between student and employment status among AYA participants (V = 0.282).

**Table 4:**
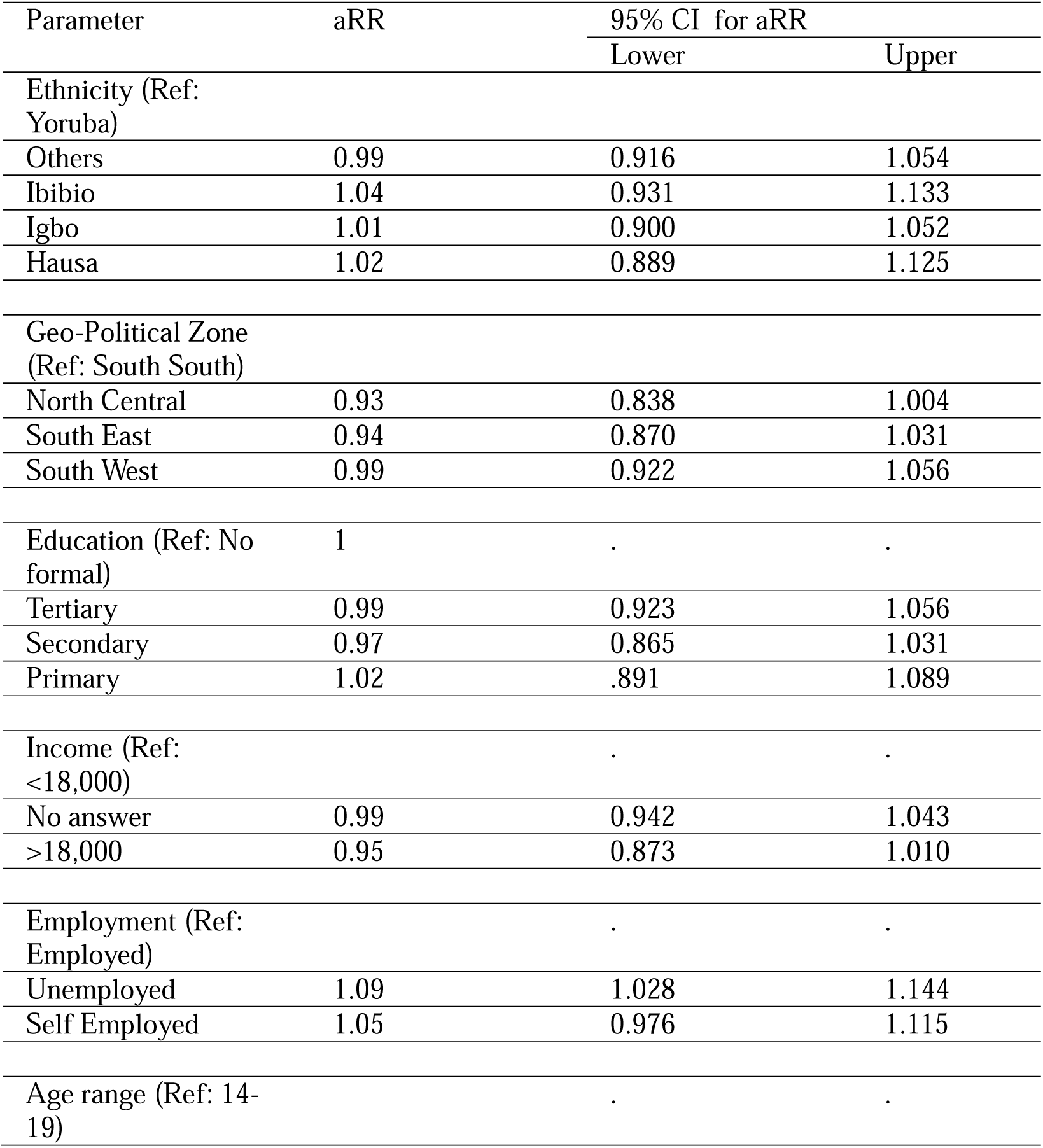
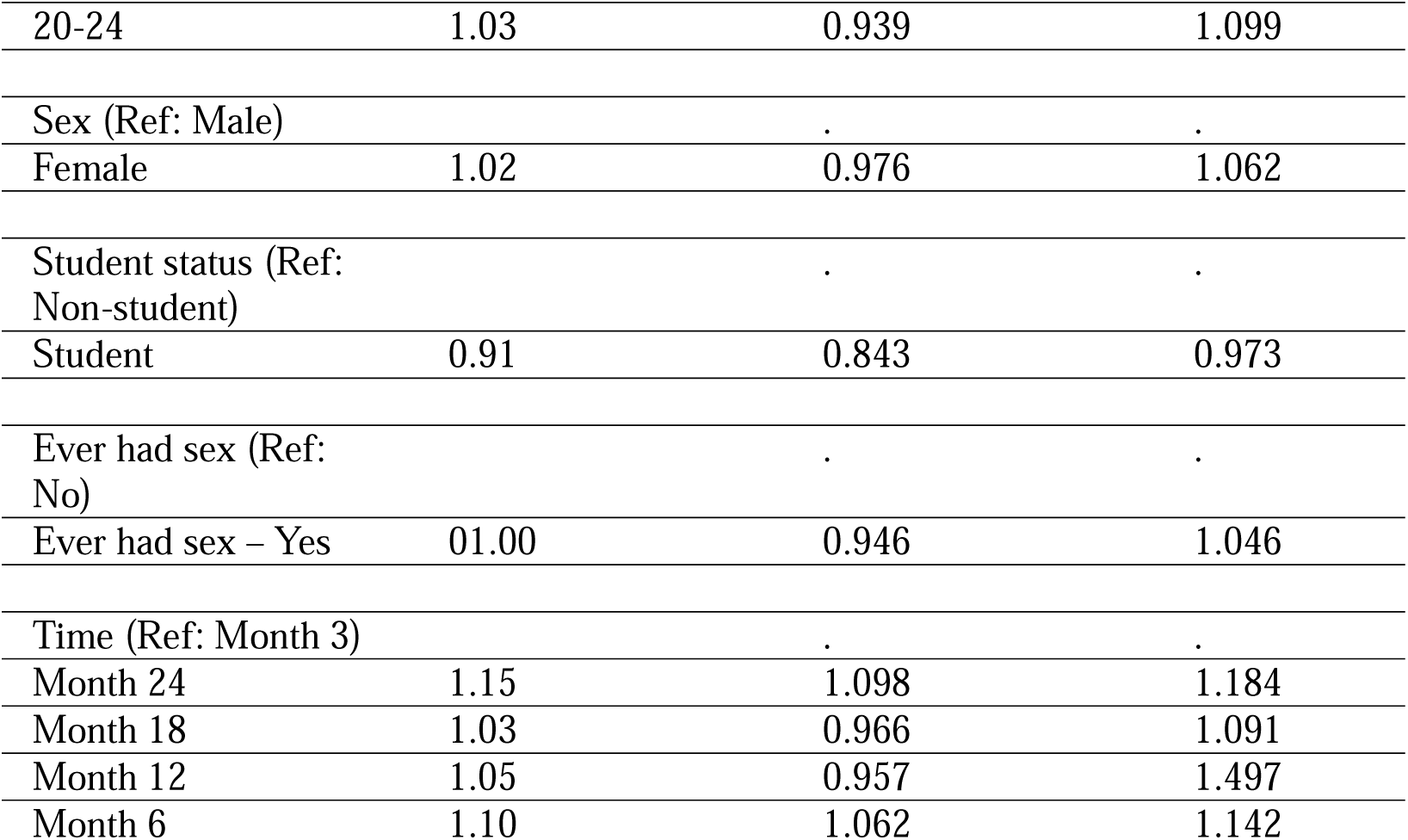
Generalized Estimating Equations Analysis of Correlates of Engagement-in HIV Prevention program.

State-level showed that literacy rate (F(4) = 15.41, p <0.001), poverty rate (F(4) = 7.33, p <0.001), and GDP per capita (F(4) = 4.41, p = 0.001) showed significant interaction with time. AYA in higher-poverty states showed significantly sustained engagement (B=-0.004, p<0.001) compared to AYA in other states. The main findings related to correlations of sustained AYA engagement were robust. Sensitivity analysis showed that participants were similar to others in terms of education, income, region, and gender, but showed a significant difference in ethnicity(p<0.001) and student status (p=0.023) (Supplement 4).

## Discussion

This study examined longitudinal relationships between socio-demographic characteristics and sustainment of AYA engagement in an HIV prevention program across 32 LGAs in Nigeria. Sustained engagement was more likely among non-students, unemployed, and people in states with a higher poverty rate. This study expands the evidence on AYA engagement in HIV programs by focusing on an under-served West African context, leveraging granular longitudinal data, and evaluating an innovative AYA-engaged intervention

We found that nearly three in four AYA sustained engagement over 24 months. The engagement rate also increased from 74.3% in month 3 to 81.5% in month 24. This finding is in contrast to a 10-year analysis among AYA where engagement decreased over time because of burnout.^29^ Also, an adolescent-centered health promotion program in Tanzania reported a sharp decline in AYA attendance in the last two contacts due to schedule conflicts with community, school, and religious activities.^30^ However, it aligns with other participatory studies suggesting that crowdsourcing, peer educators, and other participatory approaches may help to sustain AYA engagement in HIV prevention research.^9,30,31^ Hence, the engagement activities employed in the HIV prevention program might have contributed to the high engagement rate and an increase in the average number of participants meaningfully engaged in the HIV prevention program over the 24 months. This may have also contributed to the high uptake of HIV self-testing reported in the initial publication from this trial.^15^ While we acknowledge that the least engaged AYA may have dropped out of the study. The socio-demographic data of participants with more missing data were similar to those of others with completed data.

Our data showed that perception of voice or power to influence decisions in the program had the lowest mean score among all items, although higher than the cut-off for engagement (3.15/4.00). Engagement declines when there is a perception of a lack of power in decision-making.^32^ Meaningful AYA engagement requires shared decision-making to prevent tokenism.^33^ In this program, AYA led the design of the intervention, including the decision-making processes. Although the implementation was delivered by AYA research assistants, study participants had minimal involvement in decision-making. Also, the AYA who joined specific engagement activities may not have been included as participants. The monthly peer-led meetings were created to receive feedback from participants through the AYA research assistants from each site. This finding suggests the need to include leadership training for AYA research assistants to ensure shared decision-making with peers while delivering intervention. This aligns with the WHO’s framework on meaningful engagement, which emphasizes operationalization of shared decision-making among AYAs. ^34^

Our data show that students were less likely to sustain engagement compared to non-students. This could be because of competing demands such as exams, school strikes, and returning home during the holidays or festivals.^30^ Undergraduate student participants experienced a major strike action that closed all federal universities across Nigeria during the implementation period.^35^ This could have resulted in psychological stress and impacted AYA engagement.^35^

Also, our study found that unemployed participants were more likely to remain engaged in the ITEST HIV prevention program than employed participants. A study from Uganda reported that unemployed AYA were four times more likely to engage in an HIV prevention program than their employed peers, although sustainability was not described.^36^ This could be linked to the availability of unemployed AYA compared to employed AYA. Employed AYA may prefer non-working hours and weekends for engagement in the program. However, most of the AYA-friendly centers function optimally during working hours. Further analysis showed a statistically significant but weak-to-moderate association between student and employment status among AYA participants. This shows that these two variables share some overlap, but they are independent correlates of sustained engagement among AYA participants. This further strengthens the impact of the dual burden of academics and work demands in influencing sustained engagement of AYA in HIV prevention programs. Hence, emphasizing the significance of these factors as social determinants of health in AYA health programming.

AYA in states with higher poverty rates had more sustained engagement compared to AYA in other states. Poverty has been described as a barrier to health program engagement among AYA, mostly due to transportation costs.^37,38^ However, our findings suggest that meaningful engagement of AYA in community-based HIV prevention programs may mitigate this structural barrier over time. High-poverty settings often have better community cohesion, which may facilitate peer-driven engagement. This state-level finding further reinforces the individual-level GEE, where unemployed AYA showed significantly greater sustained engagement. This suggests that economic marginalization at the state or individual level may enable sustained engagement in the HIV prevention program. This could be due to the participatory approach of the intervention, where every AYA participant had equal power without discrimination based on socioeconomic status. Hence, co-creating with communities can strengthen social solidarity and collective mobilization to engage in HIV prevention despite structural or economic disadvantages.^39^ For global health program designers and policymakers, this suggests that targeting high-poverty settings for participatory HIV prevention programs may not only promote equity but also sustainability.

## Implications

This study has both policy and research implications. AYA-focused HIV programs should engage those with lived experiences from the conceptualization and design phase of the intervention through participatory activities (i.e., crowdsourcing an open call). Also, our data support investment in community-based, AYA-friendly HIV prevention programs that operate outside clinical settings and conventional working hours. This approach should be incorporated into the national HIV program to operationalize participatory AYA engagement, including shared decision-making across all phases of program design, implementation, and evaluation. This can be replicated across other programs in the health system. Program implementers should consider different strategies for the AYA population with busy schedules, such as organizing activities on weekends or non-working hours. Also, incentive structure and engagement activities should be adaptive to accommodate AYA based on their preference and characteristics, such as sexual experience, educational level, and daily schedule. For instance, while a condom may be a preferred incentive for sexually active AYA, perfume or beauty products may be preferred by non-sexually active AYA. Although, this may increase the complexity of the policy or intervention. Lastly, AYA participants need to have shared decision-making power. Peer educators or AYA organizers should be trained on AYA leadership and design an iterative feedback loop for real-time input from participants to improve and sustain engagement of participants in the program. These strategies will help implementers design equitable and sustainable AYA-friendly HIV prevention programs in low- and middle-income countries (LMICs) with adaptation to the cultural context of implementing communities.

This study also highlights research gaps. Future research should examine how AYA engagement can be sustained beyond the implementation period. Also, there is a need to explore the effectiveness of differentiated engagement strategies stratified across AYA subgroups based on sexual experience, age group, education, and employment status.

## Limitations

This study has some limitations. First, the study may be prone to selection bias because some participants did not complete all surveys. Our sensitivity analysis showed a significant difference between the studentship variable of the included and excluded data. Also, AYA participants with prior relationships with the AYA research assistants could have self-reported a higher level of engagement. Last, state-level contextual data were obtained from datasets of a single year and are assumed to be representative of conditions during the 24-month period. This was due to the unavailability of annual data for these variables and the indicators were considered to be stable during the study period.

## Conclusion

AYA engagement in the HIV prevention program was high and sustained throughout the 24 months, demonstrating the potential of AYA-led engagement strategies in sustaining HIV prevention among AYA in Nigeria. It also suggests that community-based participatory design can foster social solidarity, helping to sustain programs in socioeconomically marginalized communities. The study emphasizes the significance of a shared decision-making structure in order to achieve meaningful AYA engagement that can be sustained beyond the implementation period, and provide actionable evidence to support programmatic design, policy-making, and implementation of equitable AYA-friendly HIV prevention programs in Nigeria and across Africa.

## Data Availability

Data are available upon request.

## What is already known on this topic

- The incidence of HIV is disproportionately high among AYA in Africa.
- AYA engagement in HIV prevention programs in Africa are often transient

## What this study adds

- The mean engagement score remained consistently high throughout the 24 months (3.25±0.54 to 3.32±0.56), with 73.0% of participants demonstrating sustained engagement.
- Sustained engagement was more likely among non-students, unemployed, and more poor states.

## How this study might affect research, practice or policy **–**

- Integration of participatory, AYA-led approach into HIV prevention services promotes equity and sustainability in low-resource settings.

## Authors’ Contributions

AOB, and JDT were responsible for conceptualization, methodology, and writing the original draft. WT, TG, SD, SN, UN, NER, YB, OF, CD, TK, TO, OO, CA, PK, LA, DO, AZM, FA, TM, KM, RB, HX, GO, CO, JI, OE, JDT were responsible for reviewing and editing the draft. All authors have read and approved the final manuscript.

## Data sharing statement

Data are available upon request.

## Transparency declaration

The lead author affirms that the manuscript is an honest, accurate, and transparent account of the study being reported.

